# Widespread use of face masks in public may slow the spread of SARS CoV-2: an ecological study

**DOI:** 10.1101/2020.03.31.20048652

**Authors:** Chris Kenyon

## Abstract

**Background:** The reasons for the large differences between countries in the sizes of their SARS CoV-2 epidemics is unknown. Individual level studies have found that the use of face masks was protective for the acquisition and transmission of a range of respiratory viruses including SARS CoV-1. We hypothesized that population level usage of face masks may be negatively associated SARS CoV-2 spread.

**Methods:** At a country level, linear regression was used to assess the association between COVID-19 diagnoses per inhabitant and the national promotion of face masks in public (coded as a binary variable), controlling for the age of the COVID-19 epidemic and testing intensity.

**Results:** Eight of the 49 countries with available data advocated wearing face masks in public – China, Czechia, Hong Kong, Japan, Singapore, South Korea, Thailand and Malaysia. In multivariate analysis face mask use was negatively associated with number of COVID-19 cases/inhabitant (coef. −326, 95% CI −601- −51, P=0.021). Testing intensity was positively associated with COVID-19 cases (coef. 0.07, 95% CI 0.05-0.08, P<0.001).

**Conclusion:** Whilst these results are susceptible to residual confounding, they do provide ecological level support to the individual level studies that found face mask usage to reduce the transmission and acquisition of respiratory viral infections.

## Background

SARS CoV-2, the viral cause of COVID-19, has spread rapidly to over 190 countries [1]. There has, however, been remarkable variation in how extensively it has spread and in the national responses to this spread [1, 2]. For example, although the virus is thought to have first emerged in China, European countries such as Italy and Spain have reported roughly 30-fold higher number of infections per capita than China [1, 2]. Understanding the reasons underpinning this heterogeneity in spread is crucial to ongoing prevention efforts. The cornerstones of prevention efforts have included extensive testing, contact tracing and isolation and various forms of social distancing/quarantining [3, 4]. Whilst there have been important differences in how these were implemented in different countries, arguably the most striking difference in approach has been in the use of universal face masks in public. Whereas a number of predominantly Asian countries have promoted this practice, the World Health Organization (WHO) and most European and North American countries have not promoted this strategy [5, 6]. The head of the Chinese Center for Disease Control and Prevention has stated that the biggest mistake that Europe and the US were making in tacking COVID-19 was their failure to promote the widespread usage of face masks in public [7]. The WHO argues against universal face mask use based on a lack of evidence to support the practice, as well as a concern that using face masks will provide users with a false sense of security which may result in poorer hand hygiene and hence increased transmission [3, 5, 8]. The US Centers for Disease Control and Prevention does not recommend that people who are well wear a face mask to protect themselves from respiratory diseases, including COVID-19 [5]. In fact, the US Surgeon General stated that facemasks “are not effective in preventing (the) general public from catching coronavirus” and urged people to stop buying face masks [5].

Advocates of universal usage of face masks point to four types of evidence. Firstly, SARS CoV-1 and -2 are spread mainly through contact- and droplet-but also through airborne-transmission [6, 9]. Detailed environmental and epidemiological investigations from the large Amoy Gardens outbreak of SARS CoV-1 revealed that airborne transmission played an important role in the outbreak [10, 11]. Likewise in vitro studies demonstrate that SARS CoV-2 can be aerosolized and remain viable in the air in this form for at least 3 hours [12]. Although viral viability was not assessed, air samples from hospital rooms and toilets used by COVID-19 patients as well as from a crowded entrance to a department store tested positive for SARS CoV-2 [13]. Even if we discount the evidence of airborne transmission, face masks could play a major role in reducing droplet and possibly contact (via reduced digital-oral interactions) transmission. The second type of evidence is that from epidemiological studies showing that masks do provide this protective effect. One systematic review on the efficacy of face masks to prevent influenza, found evidence that face masks were effective in preventing the transmission to others and weaker evidence that they prevented influenza acquisition [14]. Likewise, a systematic review and metanalysis in health care workers found that mask wearing was associated with a lower incidence of clinical respiratory infections [15]. A Cochrane review of different physical measures to prevent the acquisition of respiratory viruses found face masks to be the most effective of all measures investigated - including social distancing [16]. The results were similar for studies limited to SARS CoV-1 transmission, with the authors concluding: ‘wearing a surgical mask or a N95 mask is the measure with the most consistent and comprehensive supportive evidence’ [16]. Thirdly, there is increasing evidence that a large proportion of SARS CoV-2 transmission occurs from pauci- or asymptomatic individuals. An estimated 30% of infections are truly asymptomatic and 80% mild infections [17]. Evidence is also mounting that infected individuals are infectious prior to the onset of symptoms [18]. Taken together these findings provide an explanation for why epidemiological studies have found that nondocumented infections were the infection source for 79% of documented cases in Wuhan, China [18]. In this setting limiting masks to confirmed infections is far less likely to have an impact on transmission than universal use. The key argument for universal use is thus preventing transmission and a secondary argument is preventing acquisition [7, 9].

These considerations provided the motivation for the current analysis where we assessed if there was ecological level evidence that countries that promoted face mask usage in public had a lower number of COVID-19 diagnoses per capita.

## Methods

### Dependent variable

The *cumulative number of cases of COVID-19* infection per million inhabitants on 29 March 2020 per country. This data was obtained from the European Centre for Disease Prevention and Control (ECDC) data repository: https://www.ecdc.europa.eu/en/geographical-distribution-2019-ncov-cases

### Independent variables

#### Universal face mask

A binary variable equal to 1 or 0 according to whether or not national policies promote the wearing of face masks in public regardless of symptoms. Countries were classified according to a narrative review of official national documents and other sources (STable 1).

**Table 1.**
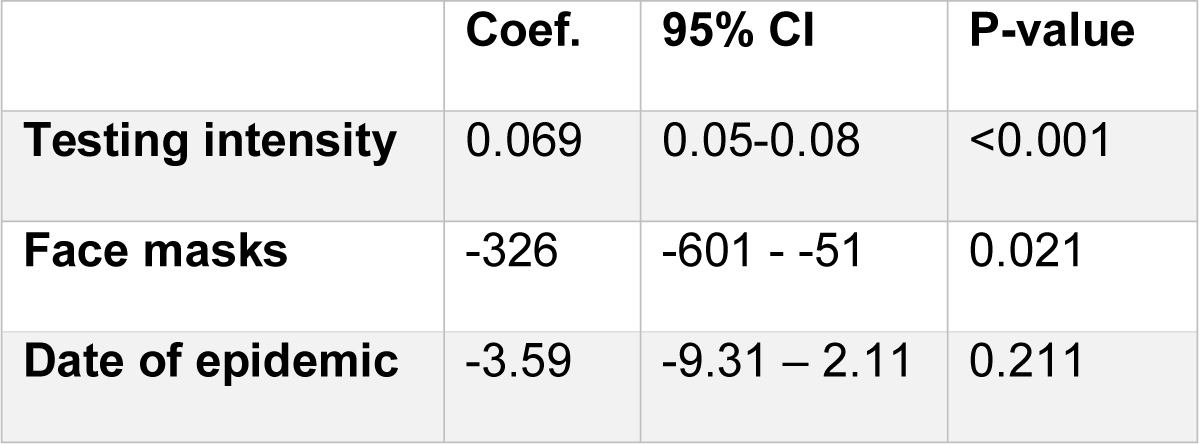
Country level, multivariate linear regression of factors associated with cumulative number of COVID-19 diagnoses per million inhabitants (N=49).

|  | **Coef.** | **95% CI** | **P-value** |
| --- | --- | --- | --- |
| **Testing intensity** | 0.069 | 0.05-0.08 | <0.001 |
| **Face masks** | -326 | -601 - -51 | 0.021 |
| **Date of epidemic** | -3.59 | -9.31 – 2.11 | 0.211 |

#### Testing intensity

Cumulative number of nucleic acid amplification SARS CoV-2 tests conducted per country per million inhabitants up till 29 March 2020. This data was extracted from the Wikipedia COVID-19 testing site on 29 March 2020: https://en.wikipedia.org/wiki/COVID-19_testing

#### Age of epidemic

The date the first case of COVID-19 was diagnosed in each country. This was measured in days after the first COVID-19 case was officially reported in China (10 January 2020). This data was obtained from the ECDC data repository on 29 March 2020: https://www.ecdc.europa.eu/en/geographical-distribution-2019-ncov-cases

## Data analysis

Linear regression was used to analyze the association between the independent and dependent variables. We controlled for the fact that SARS CoV-2 epidemic is at different stages in different countries via two methods. Firstly, the ‘age of the epidemic’ variable was included in all analyses. Secondly, we only included countries with at least 500 cumulative cases and countries whose first case was reported before 7 March 2020. Countries with missing data were dropped from the analyses. The analysis was performed in STATA version 16 (Stata Corp, College Station, Tx). Although Hong Kong is a part of China it was included as a separate data point in keeping with its population size and relative autonomy as regards public health responses. Because Czechia was the only country to introduce universal face masks late in the epidemic (18 March 2020), we repeated the analyses excluding Czechia [19, 20].

## Results

Forty-nine countries were found with complete data, epidemics older than 7 March 2020 and more that 500 cases/million inhabitants (STable 1). Large variations were evident in the number of COVID-19 cases per million inhabitants (median 158, interquartile range [IQR] 20-486), testing per million (median 1723, IQR 307-4802) and the date of the first case (median 24 February, IQR 28 January – 28 February; STable 1). Only 8 of these countries advocated wearing face masks in public – China, Czechia, Hong Kong, Japan, Singapore, South Korea, Thailand and Malaysia. These countries tended to have older epidemics. Seven of the 8 were in the group of countries with the 10 oldest epidemics (STable 1).

In multivariate analysis face mask use was negatively associated with number of COVID-19 cases (coef. -326, 95% CI -601- -51, P=0.021; Table 1). Testing intensity was positively associated with COVID-19 cases (coef. 0.07, 95% CI 0.05-0.08, P<0.001).

Repeating the analyses excluding Czechia strengthened the association between COVID-19 cases and face mask usage slightly (STable 2).

## Discussion

In this ecological study we found that countries that promoted widespread face mask usage had lower cumulative numbers of COVID-19 diagnosed after controlling for testing intensity and age of the epidemic. It is important to note that this association may be entirely explained by unmeasured confounders. For example, if countries promoting universal face masking also conducted more effective contact tracing and isolation than other countries and this was responsible for the slower spread, our study design would have falsely attributed this effect to using face masks. We did not have accurate data to control for these confounders. We did however control for testing intensity which is an important potential confounder. We also controlled for the age of the epidemic which is an obvious independent determinant of the size of the epidemic. A further limitation of our study was that we were unable to quantitate the intensity of face mask use per country. This resulted in a rather crude binary classification of face mask usage per country.

At this albeit early phase of the pandemic it is important to explore why the virus has spread more extensively in many European countries than China and neighboring countries where the outbreak appeared to have commenced. This is difficult in the absence of data that quantitates national competency in terms of the various components of successful national COVID-19 response plans. Narrative reviews of the features that resulted in the success of countries such as China, Hong Kong, Japan, Singapore and Taiwan have noted a number of common features. These include: rapid response, extensive testing, contact isolation and the widespread usage of face masks in public [4, 21].

There are a number of countries in Western European such as Italy that have conducted intensive screening, contact tracing, isolation, social distancing and widespread lockdowns and yet have amongst the largest COVID-19 epidemics in the world [2]. A striking omission from this response-list if we compare it to the responses in China and other Asian countries with lower COVID-19 incidence is that the widespread use of face masks in public was not promoted. The only European country to adopt this strategy was Czechia, and it did so at a relatively late stage in the epidemic [19, 20]. Early indications suggest that despite higher testing rates than the average for western European countries, the number of new infections is lower in Czechia [1]. Future studies will however be crucial to evaluate the impact of this intervention in Czechia and elsewhere. These studies may benefit from including data from Taiwan and Macau where use of face masks in public has been high and the cumulative number of infections has remained so low that they did not meet the 500 case threshold for inclusion in this study (Taiwan 283 and Macau 34 cumulative cases as of 29 March) [1].

It is likely that a single intervention is not sufficient to suppress the spread of COVID-19 [2]. The safest approach in the middle of this epidemic may be to introduce the full package of interventions that have been proven to work in Asian countries and then scale back according to new findings [3, 9]. Our analysis provides further evidence that this package should include widespread usage of face masks in public. Currently the only European country that can be considered to be doing this is Czechia.

## Data Availability

Data is publicly available as detailed in the Methods

## Authors’ contributions

CK conceptualized the study, was responsible for the acquisition, analysis and interpretation of data and wrote the analysis up as a manuscript.

## Ethical approval

The analysis involved a secondary analysis of public access ecological level data. As a result, no ethics approval was necessary.

## Informed consent

Not applicable

## Acknowledgements

Nil

